# Factors associated with the spatial heterogeneity of COVID-19 in France: a nationwide ecological study

**DOI:** 10.1101/2020.09.17.20196360

**Authors:** J Gaudart, J Landier, L Huiart, E Legendre, L Lehot, MK Bendiane, L Chiche, A Petitjean, E Mosnier, F Kirakoya-Samadoulougou, J Demongeot, R Piarroux, S Rebaudet

## Abstract

Like in many countries and regions, spread of the COVID-19 pandemic has exhibited important spatial heterogeneity across France, one of the most affected countries so far.

To better understand factors associated with incidence, mortality and lethality heterogeneity across the 96 administrative departments of metropolitan France, we thus conducted a geo-epidemiological analysis based on publicly available data, using hierarchical ascendant classification (HAC) on principal component analysis (PCA) of multidimensional variables, and multivariate analyses with generalized additive models (GAM).

Our results confirm a marked spatial heterogeneity of in-hospital COVID-19 incidence and mortality, following the North East – South West diffusion of the epidemic. The delay elapsed between the first COVID-19 associated death and the onset of the national lockdown on March 17^th^, 2020, appeared positively associated with in-hospital incidence, mortality and lethality. Mortality was also strongly associated with incidence. Mortality and lethality rates were significantly higher in departments with older population, but they were not significantly associated with the number of intensive-care beds available in 2018. We did not find any significant association between incidence, mortality or lethality rates and incidence of new chloroquine and hydroxychloroquine dispensations in pharmacies either, nor between COVID-19 incidence and climate, nor between economic indicators and in-hospital COVID-19 incidence or mortality.

This ecological study highlights the impact of population age structure, epidemic spread and transmission mitigation policies in COVID-19 morbidity or mortality heterogeneity.

## Introduction

Spread of the COVID-19 pandemic has exhibited important heterogeneity between countries and regions (1,2). Spatial differences between incidence and mortality rates have been associated with factors as various as: arrival time of the SARS-CoV-2 virus (3), population age structure (4), urban development and population density (5), economic level (5), health system (6), climatic and meteorological factors (7–15), anti-contagion policies and practices (16–19). In France, the first COVID-19 case was confirmed on January 24^th^ 2020. By June 10^th^, France was one of the most affected countries so far, with 150,000 cumulative confirmed cases, and nearly 30,000 associated deaths (22). While the spatial distribution of the first epidemic wave was very heterogeneous across the country (14,23,24), no study has yet analysed the underlying combination of determinants of such spatial heterogeneity. We therefore conducted an exploratory geo-epidemiological analysis based on publicly available data, to study ecological factors associated with in-hospital COVID-19 incidence, mortality and lethality (case fatality) rates across the 96 administrative departments of metropolitan France.

## Material and methods

### Brief description of the COVID-19 first epidemic wave and mitigation measures in France

In France, the first 3 patients diagnosed with SARS-CoV-2 were reported on January 24^th^, 2020, one in Bordeaux, and 2 in Paris (capital city), returning from Wuhan (China) (25–27). In February, 4 clusters were reported in other different departments, including Haute-Savoie, Oise, Morbihan and Haut-Rhin. These clusters were mainly due to secondary contaminations from people travelling from Singapore, China, Italy or Egypt, and no specific spatial trends were identified at this stage. The 2 biggest clusters were in Oise and Haut-Rhin departments. One religious event, from February 17^th^ to 21^st^ in Mulhouse city (Haut-Rhin department), brought together 2,000 to 2,500 participants from all over France and different countries (*e*.*g*., Belgium, Switzerland, Germany, Burkina Faso (28)). Most of them got contaminated and spread the virus onwards in returning home. The Oise’s cluster, near Paris, appeared in late February around a military Air base in Creil City, employing around 2,500 persons. Air Base members had been involved in the repatriation of French citizen from China on January 31^st^.

Given the accelerating number and spatial spreading of new cases and clusters, first official mitigation measures were gradually applied from March 5^th^, such as ban on gathering of more than 5,000 persons, then 1,000 persons (March 14^th^) and 100 (March 16^th^) (29). On March 15^th^, the first round of municipal elections took place but unessential services including restaurants and cafés were closed, then schools and main religious services on the following day. On March 17^th^ 2020, as France had already recorded a total of 7,730 reported cases and 175 reported death, and as incidence was doubling every other three days, a national lockdown was ordered by the government (30), and eventually extended until May 11^th^. The daily incidence peak was reached on March 31^st^, with 7,578 new confirmed cases. But considering the low level of testing capacities in France at this time of the epidemic, a model suggests that as much as 300,000 daily new infections may have occurred right before lockdown initiation (23).

### Studied parameters and data collection

Numbers of confirmed or suspected COVID-19 outpatients were not systematically recorded by the French surveillance system coordinated by *Santé Publique France*. Indeed, the majority of infectious cases, asymptomatic or even symptomatic, were not detected during the first epidemic wave, and, due to a shortage of tests, RT-PCR were reserved to in-hospital cases. Conversely, among inpatients, confirmed (by RT-PCR) or probable (according to clinical and CT-scan signs) in-hospital cases and deaths have been systematically notified. We obtained these in-hospital data at the department level from the open data portal of the French government (31), and calculated cumulative figures from March 1^st^ to April 30^th^ 2020 (Table 1), when France experienced the first epidemic wave.

**Table 1.** List of data used in the study.

| Data used in the study | Data transformation | Inclusion in GAM models |  |  | Ref |
| --- | --- | --- | --- | --- | --- |
|  |  | Incidence ratio (SIR) | Mortality ratio (SMR) | Lethality ratio (SLR) |  |
| In-hospital COVID-19 data per department between March 1<sup>st</sup> and April 30<sup>th</sup> 2020 |  |  |  |  |  |
| Cases | no | outcome | cofactor | offset | (31) |
| Deaths | no | no | outcome | outcome | (31) |
| Relative lag between the first COVID-19 associated death and the March 17<sup>th</sup> lockdown | no | cofactor | cofactor | cofactor |  |
| Total population | no | offset | offset |  | (36) |
| Population age structure estimated in 2020 per department | classification | cofactor | cofactor | cofactor |  |
| Population per 5-year and sex strata |  |  |  |  | (36) |
| Intensive-care capacity | no | no | cofactor | cofactor |  |
| Number of intensive-care beds in 2018 |  |  |  |  | (37) |
| Number of new chloroquine and hydroxychloroquine dispensations in pharmacies from January 1<sup>st</sup> to April 16<sup>th</sup> 2020 | no | cofactor | cofactor | cofactor | (38) |
| Baseline population health and healthcare services | classification | cofactor | cofactor | cofactor |  |
| Basal number of total deaths per department between March 1<sup>st</sup> and April 30<sup>th</sup> 2018 and 2019 |  |  |  |  | (40) |
| Availability of healthcare resources: densities of medical doctors, general practitioners, nurses, pharmacies and hospital beds per inhabitants |  |  |  |  | (41) |
| Subsidized medical or dependency insurance: proportion of the population covered by Universal Health Insurance (CMU), proportion of the population receiving home nursing care, proportion of the elderly population recipient of the autonomy allowance (APA) at home |  |  |  |  | (41) |
| Ageing index |  |  |  |  | (36) |
| Incidence rate of hospital stays in endocrinology, cardiology, pneumology and medicine wards |  |  |  |  | (42) |
| Economic indicators | classification | cofactor | no | no |  |
| Poverty ratio |  |  |  |  | (41) |
| Median standard of living |  |  |  |  | (41) |
| Unemployment ratio |  |  |  |  | (41) |
| Housing allowance ratio |  |  |  |  | (41) |
| Earned income supplement (RSA) ratio |  |  |  |  | (41) |
| Average amount of the autonomy allowance (APA) |  |  |  |  | (41) |
| Social assistance ratio |  |  |  |  | (41) |
| **Urbanization** | classification | cofactor | no | no |  |
| Proportion of the population living in metropolitan cities |  |  |  |  | (41) |
| Proportion of the population living in multipolar cities |  |  |  |  | (41) |
| Proportion of the population living in remote communes |  |  |  |  | (41) |
| Proportion of the population living in other communes |  |  |  |  | (41) |
| Roads density outside urban areas |  |  |  |  | (41) |
| Surface area |  |  |  |  |  |
| **Climate (% of surface area)** | classification | cofactor | no | no |  |
| Mountain; semi-continental and sub-montane; modified oceanic of centre and north plains; altered oceanic; franc oceanic; altered Mediterranean; south-west basin; franc Mediterranean |  |  |  |  | (43) |
| **Spatial** | spline | cofactor | cofactor | cofactor |  |
| Centroid coordinates |  |  |  |  | (44) |

Outcome in generalized additive models : SIR, standardized incidence ratio; SMR, standardized mortality ratio; SLR, standardized lethality ratio

We also calculated the relative lag between the first COVID-19-associated confirmed death and the March 17^th^ lockdown for each department in order to account for the temporal progression of the epidemic wave across the country and the effect of the national lockdown. As male sex and older age are associated with increased severity (32–35), we extracted the population age and sex structure estimated in 2020 for each department, from the French national statistics institute (*Institut national de la statistique et des études économiques* INSEE) (36). Considering the importance of intensive-care capacities in COVID-19 care, we obtained the number of intensive-care beds per department in 2018 from the Ministry of health website (37). During the early phase of the epidemic, very limited evidence existed regarding treatments with potential antiviral properties. Among the many treatments proposed, the French national drug agency (*Agence nationale de sécurité du médicament* et *des produits de santé*, ANSM) and the French national health insurance fund (*Caisse nationale d’assurance maladie*, CNAM) observed a surge in new chloroquine and hydroxychloroquine (CQ/HCQ) dispensations in pharmacies during the epidemic (38). As the role of CQ/HCQ consumption in the spatial heterogeneity of COVID-19 incidence and mortality rates have been evoked (20), we extracted the number of new dispensations at department-level between January and April 16^th^ 2020 from an official report (38). We also considered additional indicators describing department-specific baseline population health and healthcare services, as the impact of structural health determinants and comorbidity have been associated to severity (39) (Table 1): baseline number of total deaths per department between March 1^st^ and April 30^th^ 2018 and 2019 (40); indicators related to the availability of healthcare resources (41); indicators of subsidized medical or dependency insurance (41); incidence ratio of hospital stays in several type of wards (endocrinology, cardiology, pneumology) (42). We gathered several economic indicators at department-level (Table 1) (41). We also obtained urbanization-describing indicators (Table 1) (41). Finally, we characterized departments according to 8 climate profiles (Table 1) (43). To take into account spatial autocorrelation, we extracted the centroid coordinates of each department from a shapefile (44).

### Statistical analyses

We calculated in-hospital COVID-19 incidence, mortality and lethality (or case-fatality) rates for each administrative department of metropolitan France. We mapped these indicators and the deviations from their mathematical expectation, and estimated spatial autocorrelation using Moran’s I statistic for areal data (Queen criterion of contiguity) (45).

In order to analyse factors associated with epidemiologic indicators, and avoid the curse of dimensionality and multiple collinearities, we initially summarized a multidimensional factor using a non-supervised classification technique: hierarchical ascendant classification (HAC) on principal component analysis (PCA) (46,47). To classify department into similar age–pyramid profiles, we performed a PCA on the relative composition of the five-year age and sex population groups, and then included the coordinates of each variable in the first 25 principal components, in a HAC. Similarly, we used HAC based on PCA coordinates to classify department population health and healthcare services, economy, climate and urbanization, based on corresponding indicators (Table 1).

We mapped the resulting classifications at the department-level, as well as time to lockdown, intensive-care beds, CQ/HCQ dispensation incidence.

In order to assess the relationships between these factors and in-hospital COVID-19 incidence, mortality and lethality rates, we used generalized additive models (GAM) with a negative binomial regression to take into account over-dispersion. A Gaussian kriging smoother (with a power exponential covariance function) based on geographical coordinates of each department centroid (*s*(*Lon, Lat*)) was used to take spatial autocorrelation into account, as previously described (48,49). The log population was used as an offset to estimate standardized incidence ratios (SIRs, Eq. 1) and standardized mortality ratios (SMR, Eq. 2), and the log cases to estimate standardized lethality ratios (SLRs, Eq. 3).

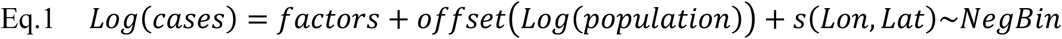

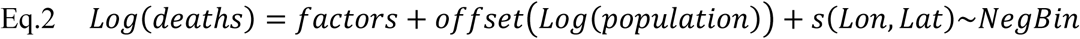

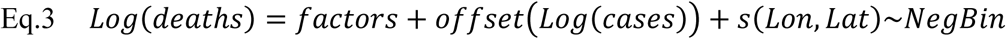

For each outcome, analysed cofactors are listed in Table 1. Briefly, to model department in-hospital incidence rate, we considered progression of the epidemic wave and all risk factors except intensive care facilities. To model in-hospital mortality rate, we considered COVID-19 in-hospital cases, factors possibly associated with case severity (population age structure, CQ/HCQ dispensation), factors associated with case management capacities and preparedness (progression of the epidemic wave, intensive-care capacity and population health and healthcare services), but not factors directly associated with incidence (economy, urbanization and climate). We considered similar factors to model in-hospital lethality, except that cases were used as an offset, to estimate standardized lethality ratios. We first separately analysed each factor with univariate models. Multivariate generalized additive models then included variables for which p-values were less than 0.25 (50), or which appeared important from a public health perspective, with the help of directed acyclic graphs (DAG) (51), such as population age structure or population health and healthcare services. In the multivariate mortality model, we considered the interaction between in-hospital cases and relative lag between the first COVID-19 associated death and lockdown. Finally, we mapped Pearson residuals of multivariate models to highlight outliers. Spatial adjustments were verified by Moran I statistics on Pearson residuals.

The level of significance was fixed at *α* = 0.05. All analyses and maps were performed using the software program R v4.0.0 (The R Foundation for Statistical Computing, Vienna, Austria), and the {*tidyverse*}, {*mgcv*}, {*FactoMineR*}, {*rgdal*}, {*spdep*} and {*dagitty*} packages.

### Ethics approval

This study did not require any research ethics approval, as analyses exclusively included publicly available, anonymized aggregated data.

## Results

### Incidence, mortality and case-fatality rates of in-hospital COVID-19

From March 1^st^ to April 30^th^ 2020, hospitals in metropolitan France notified a total of 94,935 COVID-19 cases, including 15,790 cases admitted in intensive-care and 15,435 deaths. This corresponded to a global cumulative in-hospital incidence rate of 146.1 cases /100,000 inhabitants, ranging from 19.0 to 456.7 cases /100,000 inhab. across the 96 administrative departments (Fig. 1). The global cumulative in-hospital mortality rate was 23.8 deaths /100,000 inhab. (from 1.1 to 107.0 deaths /100,000 inhab.). Both indicators exhibited a marked heterogeneity following a north-east to south-west gradient (Fig. 1), with high and significant spatial autocorrelations (both Moran’s I statistics, 0.68; p-values <0.001) related to the diffusion of the epidemic. Highest incidence and mortality rates were observed in Territoire-de-Belfort and Haut-Rhin with respectively 456.7 and 456.1 cases /100,000 inhab., and 107 and 95.8 deaths /100,000 inhab.. In Paris department, incidence and mortality rates were 378.4 cases and 68.7 deaths /100,000 inhab., respectively. Incidence and mortality rates in Bouches-du-Rhône (department including Marseille the 2^nd^ largest French city) were 186.9 cases and 20.2 deaths /100,000 inhab., respectively, which were higher than in surrounding departments. The global cumulative in-hospital lethality rate in metropolitan France was 16.3%. The lethality rate exhibited a marked heterogeneity (from 4.7% to 25.2% across departments, resp. Ariège and Vosges) (Fig. 1), but with a lower spatial autocorrelation (Moran’s I statistic, 0.32; p-values <0.005).

**Fig. 1.**
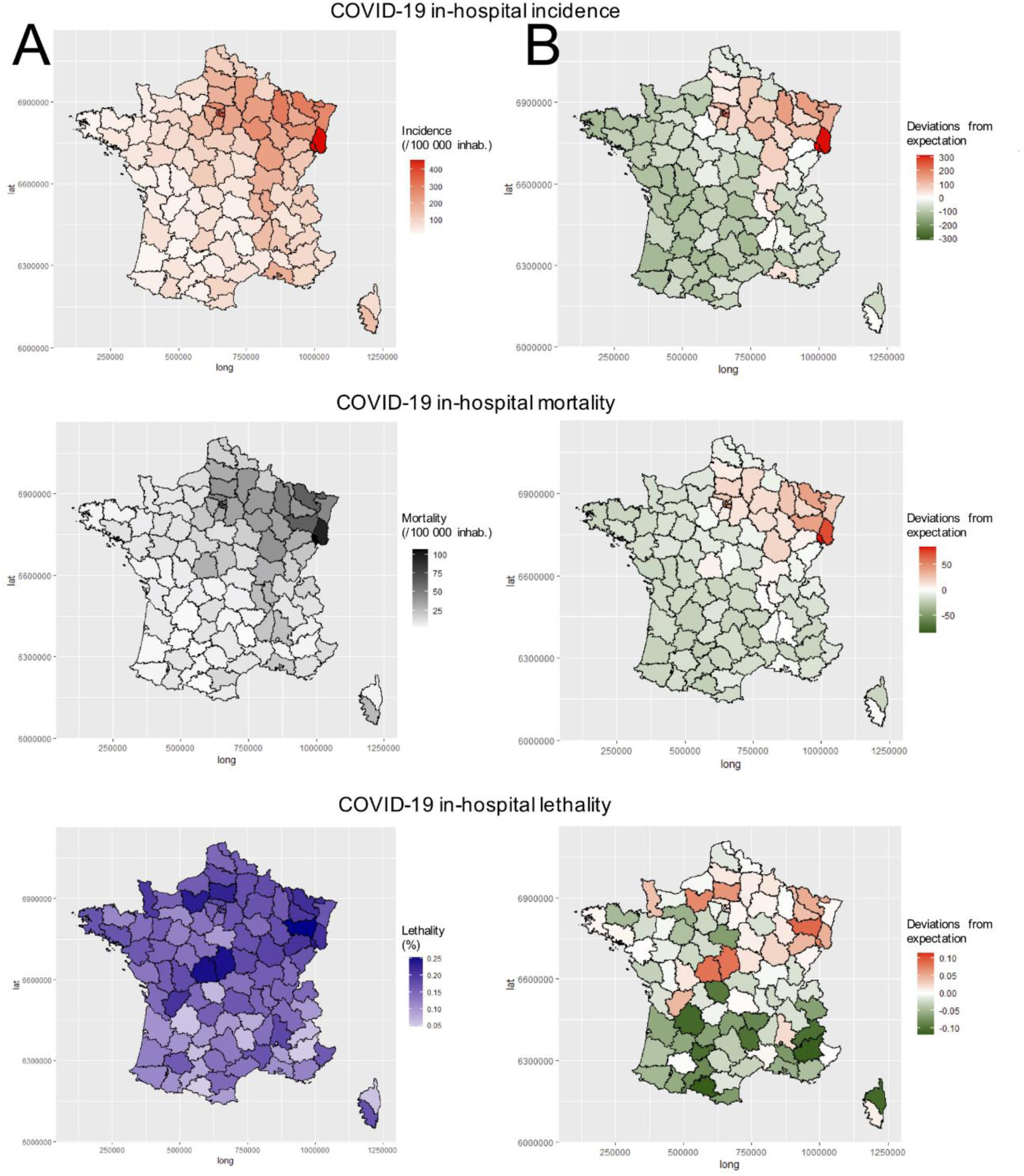
(A) Cumulative in-hospital incidence, mortality and lethality rates in the 96 departments of metropolitan France; and (B) deviations from mathematical expectation.

### Description and classification of covariates

In median across the 96 departments, the first COVID-19 associated death occurred 3 days after the March 17^th^ lockdown, *i*.*e*. median relative lag of -3 days (Fig. 2A), with a maximum 19 days in Oise, and a minimum -44 days in Lozère. This lag illustrated the temporal progression of the epidemic wave across the country along a north-east to south-west axis.

**Fig. 2.**
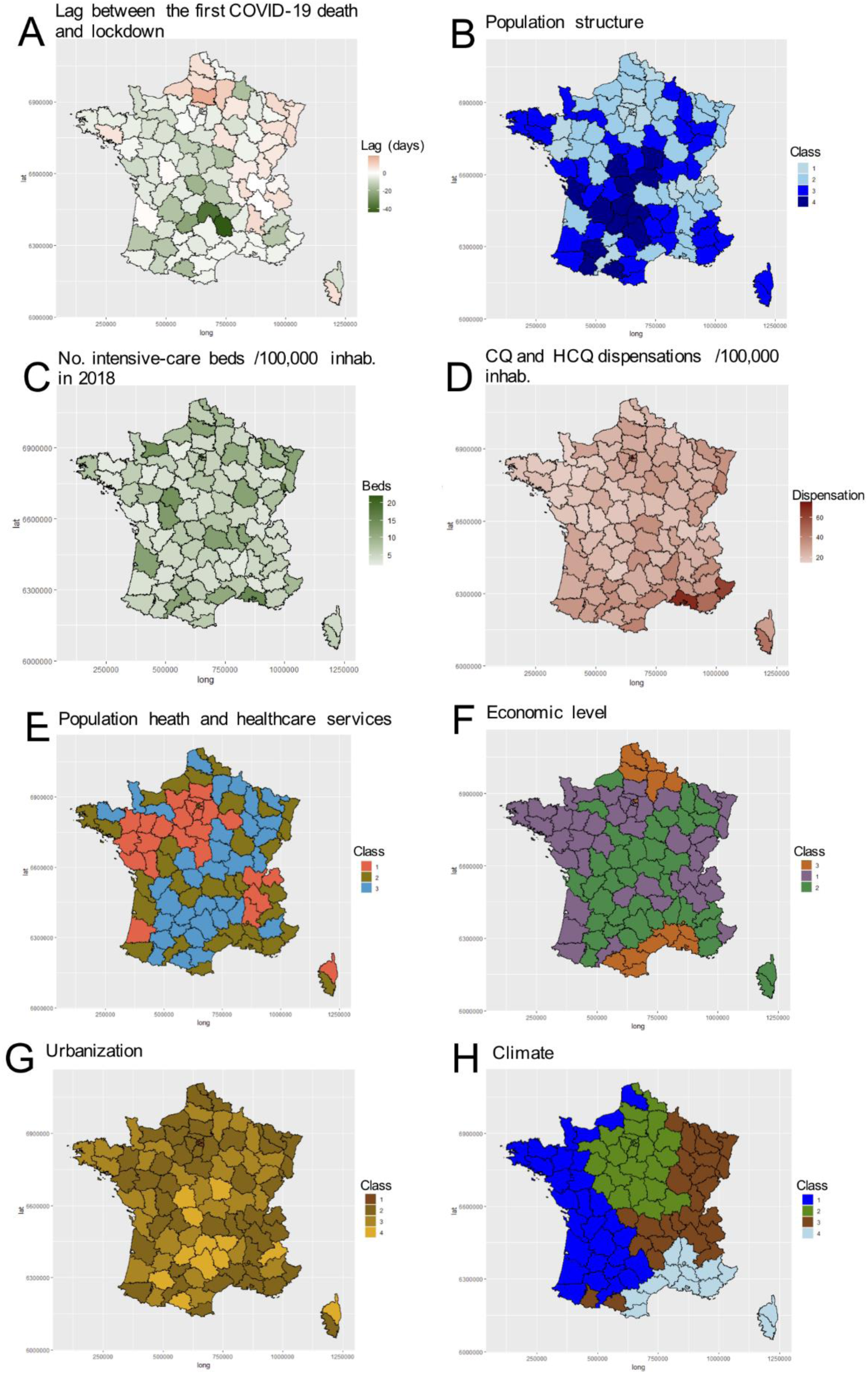
Maps of department covariates. Classes are defined in the main text and in Table 2.

HAC based on PCA coordinates from age pyramids classified departments into four classes (Fig. 2B): (Class 1) a majority of 25-45 years-old inhabitants; (Class 2) high proportion of <25 years-old inhabitants; (Class 3) high proportion of 50-85 years-old inhabitants; and (Class 4) high proportion of >85 years-old inhabitants, whatever gender characteristics. The global density of intensive-care beds in 2018 in France was 8.1 /100,000 inhab., ranging from 2.0 in Eure, to 21.9 in Paris (Fig. 2C). The global new CQ/HCQ dispensations in French pharmacies was 32.0 /100,000 inhab. from January 1^st^ to April 16^th^ 2020, and ranged from 14.4 in Mayenne, to 68.7 in Bouches-du-Rhône and 75.5 in Paris (Fig. 2D). PCA and HAC classified departments into three classes of population health and healthcare services (Fig. 2E): (Class 1) high proportion of the population receiving at home health assistance; (Class 2) high health professional density; (Class 3) high proportion of hospital stays (endocrinology, pneumology, cardiology). Classification on economic indicators classified departments into three classes (Fig. 2F): (Class 1) high median standard of living; (Class 2) high rate of social assistance; (Class 3) high poverty and unemployment ratios. Classification on urbanization indicators classified departments into four classes (Fig. 2G): (Class 1) very high proportion of the population living in metropolitan cities and high roads density (Paris and surrounding metropolitan departments); (Class 2) high proportion of the population living in metropolitan cities and lower roads density; (Class 3) high proportion of the population living in multipolar cities; (Class 4) high proportion of the population living in remote communes. Finally, classification on climate indicators identified four department classes (Fig. 2H): (Class 1) central plains with modified oceanic climate; (Class 2) departments with oceanic or south-west basin climate; (Class 3) departments with semi-continental, sub-montane or mountain climate; and (Class 4) departments with altered or frank Mediterranean climate.

**Table 2.**
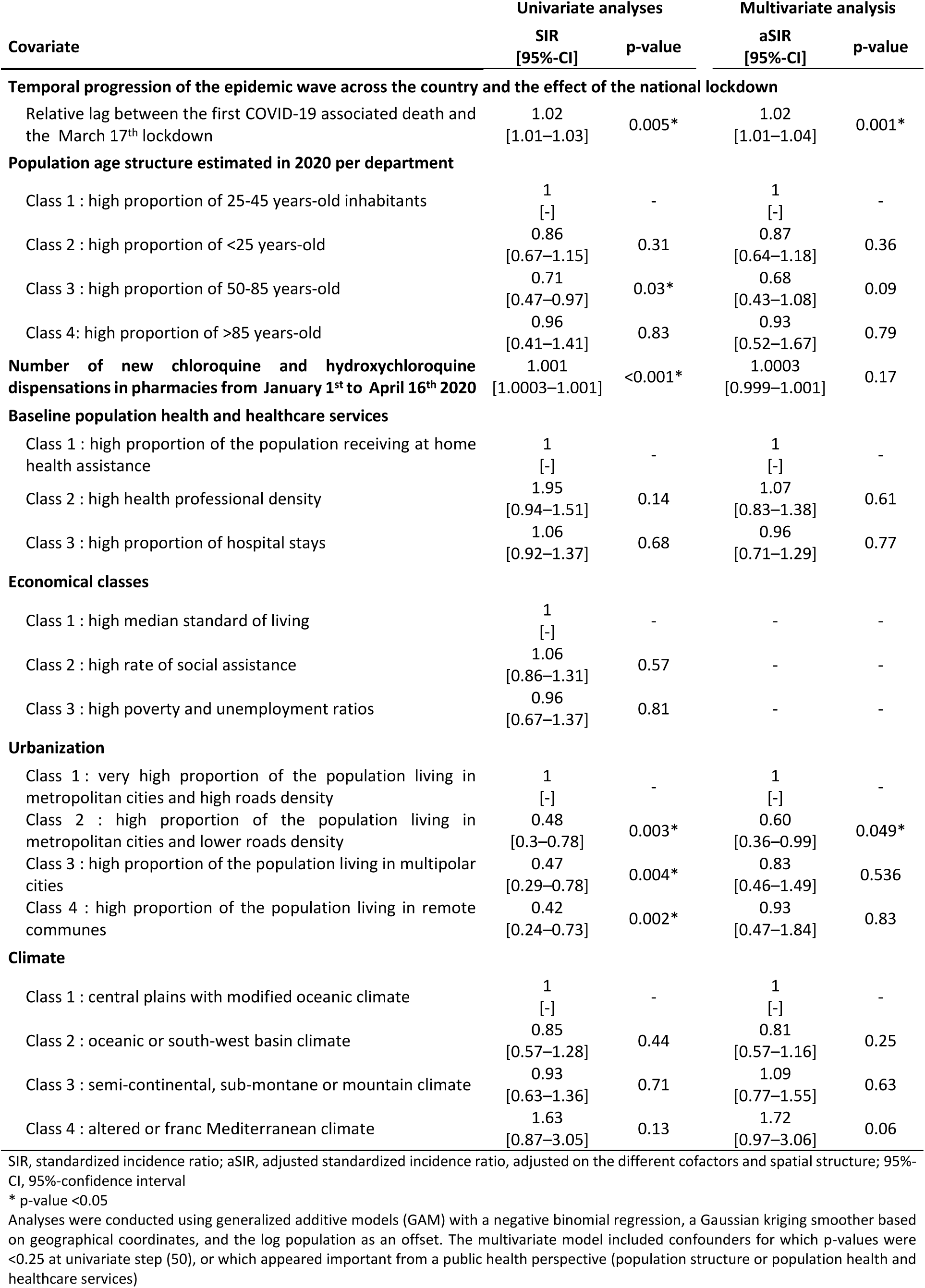
Factors associated with in-hospital COVID-19 incidence rate at the department level in metropolitan France.

### Factors associated with in-hospital COVID-19 incidence rate at the department level

In univariate analyses, COVID-19 incidence rate from March 1^st^ to April 30^th^ at the department level was significantly associated with the lag between the first death and lockdown, the population age structure, new CQ/HCQ dispensations, and urbanization (Table 2).

In the final multivariate model, early arrival of the epidemic wave was associated with higher cumulative COVID-19 incidence rates (adjusted standardized incidence ratio for each day between the first death and lockdown, aSIR = 1.023 [95%-confidence interval, 1.009–1.037]; p-value = 0.001) (Table 2). Compared to departments of Paris and surrounding metropolis, (Class 1 of urbanization), departments with a high proportion of the population living in metropolitan cities and lower roads density (Class 2) exhibited a significantly lower COVID-19 incidence rate (aSIR = 0.60 [0.36–0.99]; p-value = 0.049). In this model, population age structure, CQ/HCQ dispensations, population health and healthcare services, and climate were not significantly associated with in-hospital COVID-19 incidence rates (Table 2).

This multivariate model significantly explained 87.7% of the in-hospital COVID-19 incidence deviance. Model Pearson residuals, which show no significant spatial correlation (Moran’s I statistics, -0.13; p-value = 0.96), are mapped in Fig 3A. None of most affected departments exhibited important model residuals. Of interest, the model particularly exhibited minimal residuals in Paris and Bouches-du-Rhône (Fig 3A).

**Fig. 3.**
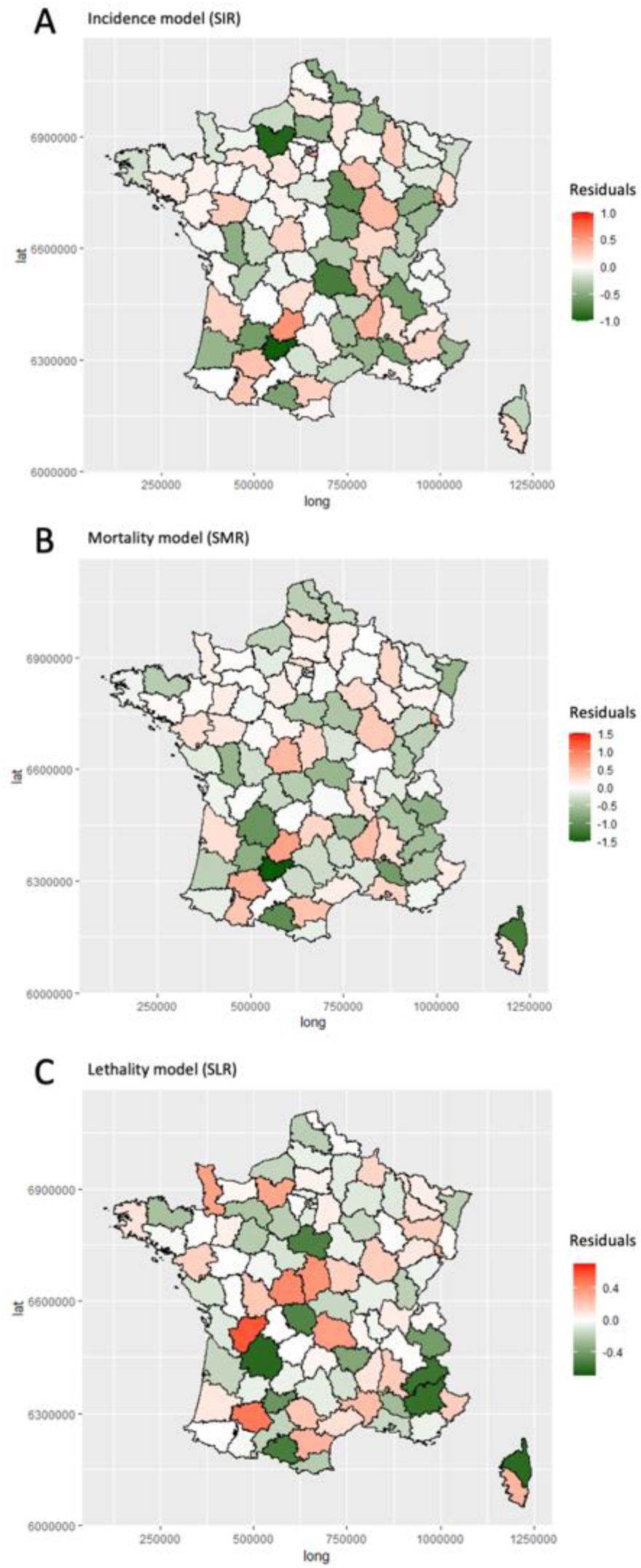
Maps of (A) incidence, (B) mortality and (C) lethality models residuals.

### Factors associated with in-hospital COVID-19 mortality rate at the department level

In univariate analyses, COVID-19 mortality rate at the department level appeared significantly and positively associated with the lag between the first death and lockdown, the cumulative number of in-hospital COVID-19 cases from March 1^st^ to April 30^th^, 2020, the intensive-care capacity, and new CQ/HCQ dispensations (Table 3).

**Table 3.** Factors associated with in-hospital COVID-19 mortality rate at the department level in metropolitan France.

| Covariate | Univariate analyses |  | Multivariate analysis |  |
| --- | --- | --- | --- | --- |
|  | SMR<br>[95%-CI] | p-value | aSMR<br>[95%-CI] | p-value |
| **In-hospital COVID-19 incidence** |  |  |  |  |
| Number of in-hospital cases accumulated from March 1<sup>st</sup> to April 30<sup>th</sup>, 2020 | 1.0002<br>[1.0001–1.0003] | <0.001* | 1.0004<br>[1.0002–1.0006] | <0.001* |
| **Temporal progression of the epidemic wave across the country and the effect of the national lockdown** |  |  |  |  |
| Relative lag between the first COVID-19 associated death and the March 17<sup>th</sup> lockdown | 1.03<br>[1.02–1.05] | <0.001* | 1.04<br>[1.02–1.06] | <0.001* |
| **Population age structure estimated in 2020 per department** |  |  |  |  |
| Class 1: high proportion of 25-45 years-old inhabitants | 1 [-] | - | 1 [-] | - |
| Class 2: high proportion of <25 years-old | 0.88<br>[0.62–1.24] | 0.45 | 1.27<br>[0.91–1.78] | 0.16 |
| Class 3: high proportion of 50-85 years-old | 0.74<br>[0.51–1.07] | 0.11 | 1.41<br>[0.90–2.19] | 0.13 |
| Class 4: high proportion of >85 years-old | 0.92<br>[0.57–1.49] | 0.73 | 2.08<br>[1.17–3.72] | 0.013* |
| **Intensive-care capacity** |  |  |  |  |
| Number of intensive-care beds in 2018 | 1.002<br>[1.0004–1.003] | 0.01* | 0.999<br>[0.995–1.003] | 0.52 |
| **Number of new chloroquine and hydroxychloroquine dispensations in pharmacies from January 1<sup>st</sup> to April 16<sup>th</sup> 2020** | 1.001<br>[1.0002–1.001] | 0.004* | 0.9994<br>[0.998–1.001] | 0.32 |
| **Baseline population health and healthcare services** |  |  |  |  |
| Class 1: high proportion of the population receiving at home health assistance | 1 [-] | - | 1 [-] | - |
| Class 2: high health professional density | 1.16<br>[0.88–1.53] | 0.29 | 0.92<br>[0.68–1.25] | 0.61 |
| Class 3: high proportion of hospital stays | 0.97<br>[0.71–1.32] | 0.84 | 0.83<br>[0.59–1.16] | 0.27 |

SMR, standardized mortality ratio; aSMR, adjusted standardized mortality ratio, adjusted on the different cofactors, spatial structure, and the interaction between COVID-19 cases and temporal progression of the epidemic wave (p=0.045); 95%-CI, 95%-confidence interval
\* p-value <0.05
Analyses were conducted using generalized additive models (GAM) with a negative binomial regression, a Gaussian kriging smoother based on geographical coordinates, and the log population as an offset. The multivariate model included confounders for which p-values were <0.25 at univariate step (50), or which appeared important from a public health perspective (population structure or population health and healthcare services)

In the final multivariate model, early arrival of the epidemic wave was associated with higher in-hospital COVID-19 mortality rates (aSMR for each day between the first death and lockdown = 1.04 [1.02–1.06]; p-value < 0.001) and with in-hospital COVID-19 case number (aSMR for each case = 1.0004 [1.0002–1.0006]; p-value < 0.001), even after taking into account a significant interaction between both factors (p-value = 0.045) (Table 3). Compared with departments with a high proportion of 25-45 years-old inhabitants (Class 1 of population age structure), departments with a high proportion of >85 years-old inhabitants (Class 4) exhibited a significantly much higher in-hospital COVID-19 mortality rate (aSMR = 2.08 [1.17–3.72]; p-value = 0.049) (Table 3). In this model, CQ/HCQ dispensations, intensive-care capacity, and population health and healthcare services were not significantly associated with in-hospital COVID-19 mortality rates (Table 3).

This multivariate model significantly explained 86.1% of the in-hospital COVID-19 mortality deviance. Model Pearson residuals, which show no significant spatial correlation (Moran’s I statistics, -0.14; p-value = 0.98), are mapped in Fig 3B. Residuals in Bouches-du-Rhône appeared higher than in surrounding departments, meaning that observed mortality was higher than the predicted mortality (Fig 3B).

### Factors associated with in-hospital COVID-19 lethality rates at the department level

In univariate analyses, COVID-19 lethality rate at the department level appeared only significantly associated with the lag between the first death and lockdown (Table 4).

**Table 4.**
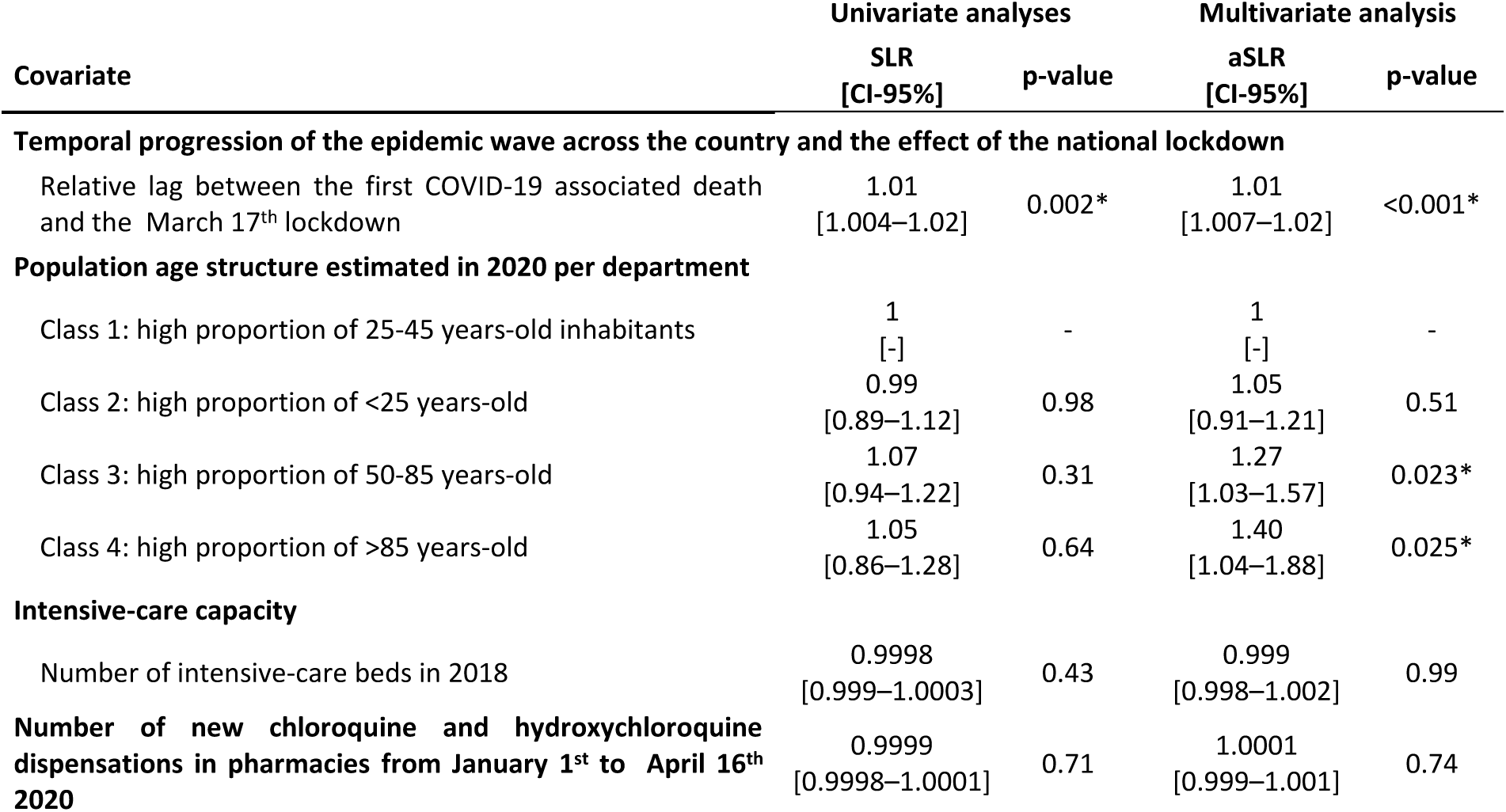
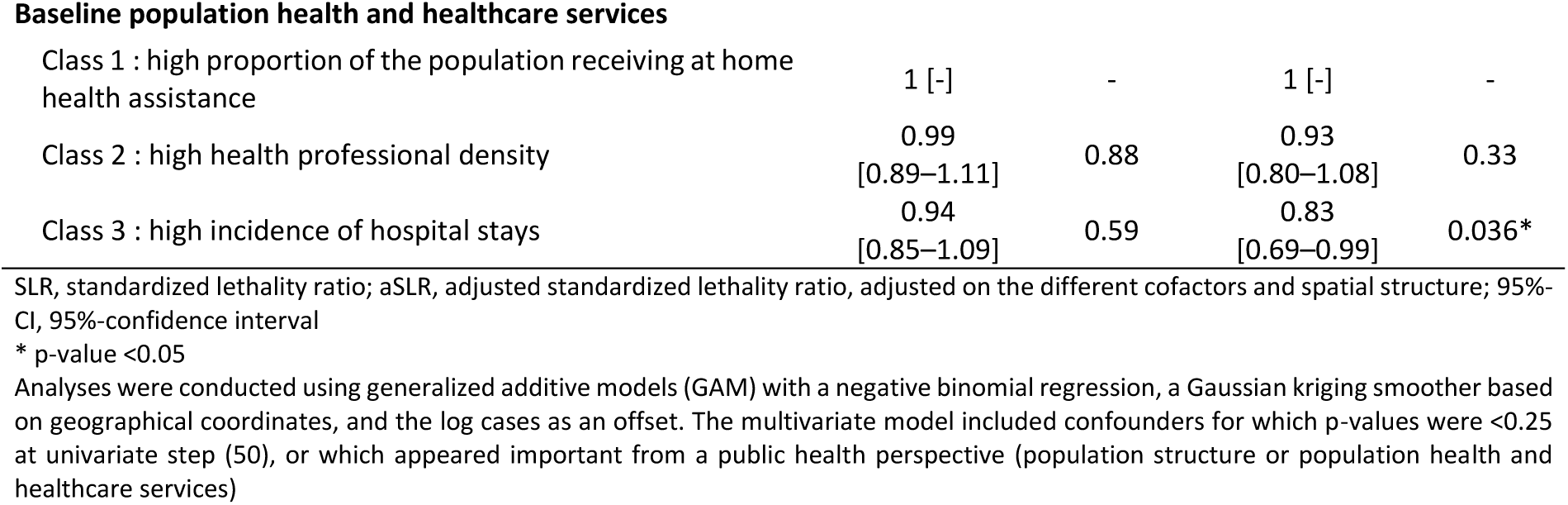
Factors associated with in-hospital COVID-19 lethality rates at the department level in metropolitan France.

| Covariate | Univariate analyses |  | Multivariate analysis |  |
| --- | --- | --- | --- | --- |
|  | SLR<br>[CI-95%] | p-value | aSLR<br>[CI-95%] | p-value |
| **Temporal progression of the epidemic wave across the country and the effect of the national lockdown** |  |  |  |  |
| Relative lag between the first COVID-19 associated death and the March 17<sup>th</sup> lockdown | 1.01<br>[1.004–1.02] | 0.002* | 1.01<br>[1.007–1.02] | <0.001* |
| **Population age structure estimated in 2020 per department** |  |  |  |  |
| Class 1: high proportion of 25-45 years-old inhabitants | 1<br>[-] | - | 1<br>[-] | - |
| Class 2: high proportion of <25 years-old | 0.99<br>[0.89–1.12] | 0.98 | 1.05<br>[0.91–1.21] | 0.51 |
| Class 3: high proportion of 50-85 years-old | 1.07<br>[0.94–1.22] | 0.31 | 1.27<br>[1.03–1.57] | 0.023* |
| Class 4: high proportion of >85 years-old | 1.05<br>[0.86–1.28] | 0.64 | 1.40<br>[1.04–1.88] | 0.025* |
| **Intensive-care capacity** |  |  |  |  |
| Number of intensive-care beds in 2018 | 0.9998<br>[0.999–1.0003] | 0.43 | 0.999<br>[0.998–1.002] | 0.99 |
| **Number of new chloroquine and hydroxychloroquine dispensations in pharmacies from January 1<sup>st</sup> to April 16<sup>th</sup> 2020** | 0.9999<br>[0.9998–1.0001] | 0.71 | 1.0001<br>[0.999–1.001] | 0.74 |
| Baseline population health and healthcare services |  |  |  |  |
| Class 1 : high proportion of the population receiving at home health assistance | 1 [-] | - | 1 [-] | - |
| Class 2 : high health professional density | 0.99<br>[0.89–1.11] | 0.88 | 0.93<br>[0.80–1.08] | 0.33 |
| Class 3 : high incidence of hospital stays | 0.94<br>[0.85–1.09] | 0.59 | 0.83<br>[0.69–0.99] | 0.036* |

SLR, standardized lethality ratio; aSLR, adjusted standardized lethality ratio, adjusted on the different cofactors and spatial structure; 95%-CI, 95%-confidence interval
\* p-value <0.05
Analyses were conducted using generalized additive models (GAM) with a negative binomial regression, a Gaussian kriging smoother based on geographical coordinates, and the log cases as an offset. The multivariate model included confounders for which p-values were <0.25 at univariate step (50), or which appeared important from a public health perspective (population structure or population health and healthcare services)

In the final multivariate model, early arrival of the epidemic wave was associated with higher in-hospital COVID-19 lethality rates (aSLR for each day between the first death and lockdown = 1.01 [1.01–1.02]; p-value < 0.001) (Table 4). Compared with departments with high proportion of 25-45 years-old inhabitants (Class 1 of population structure), departments with high proportion of 50-85 years-old inhabitants (Class 3) and with high proportion of >85 years-old inhabitants (Class 4) exhibited a significantly higher COVID-19 lethality rate (Table 4). Compared with departments with high proportion of the population receiving at home health assistance (Class 1 of population health and healthcare services), departments with high incidence of hospital stays (Class 3) exhibited a significantly higher COVID-19 lethality rate (Table 4).

This multivariate model significantly explained 53.1% of the in-hospital COVID-19 lethality deviance. Model Pearson residuals, which show no significant spatial correlation (Moran’s I statistics, -0.13; p-value = 0.96), are mapped in Fig 3C. Residuals appeared higher in less affected departments. In Bouches-du-Rhône, the model slightly overestimated lethality rate (Fig 3C).

## Discussion

Our geo-epidemiological study highlighted a marked spatial heterogeneity of in-hospital COVID-19 incidence and mortality across departments of metropolitan France, following the North East – South West diffusion of the epidemic before the national lockdown was declared on March 17^th^, 2020. Indeed, following initial sporadic cases and limited clusters in several departments (52), the COVID-19 epidemic wave began in north-eastern France (Grand-Est) and rapidly spread towards departments of more northern central regions (Ile-de-France and Haut-de-France). Hampered by the lockdown, the epidemic reached the rest of the country more slowly. In these other departments, the first deaths generally occurred after lockdown implementation, except for specific departments such as Corse-du-Sud (Corsica Island) and Morbihan (Brittany). In the south-eastern region, the most densely populated department (Bouche-du-Rhône including Marseille, the second largest French city) was more affected than the surrounding ones, in terms of incidence and mortality rates. In-hospital lethality was also heterogeneous but exhibited a less clear, albeit significant, spatial pattern, probably due to multifactorial local contexts.

After classification of multidimensional variables and multivariate analyses, the delay elapsed between the first COVID-19 associated death and the onset of lockdown appeared positively associated with in-hospital incidence, mortality and lethality, which respectively increased by 2.3% [0.9–3.7%], 4.2% [2.3–6.1%] and 1.4% [0.7–2.2%] per day of delay. In other words, morbidity and mortality were significantly lower in departments where the March 17^th^ general lockdown date caught the epidemic at earlier stages of spread. This strongly suggests that lockdown was an effective way to control the diffusion of the epidemic wave across the country, in spite of high social and economic costs. This effect of the lockdown strategy is in line with other publications based on modelling or observational studies. Among them, Salje (23) and Di Domenico (53) estimated that the lockdown resulted in a 77% and 81% reduction in transmission in France and Ile-de-France respectively; based on databases across 149 countries, Islam estimated that early implementation of lockdown was associated with a larger incidence rate reduction (54).

As expected, mortality was also strongly associated with the incidence of in-hospital COVID-19 cases, even after adjusting for the interaction between incidence and the relative lag to lockdown. Besides, the number of intensive-care beds available in 2018 was not significantly associated with mortality and lethality. This suggests that hospitals could manage to scale up their intensive-care capacity or organize medical evacuations to less affected departments when necessary. This scale-up capacity may have been higher in departments with important hospitals, as indicated by the significant negative association between lethality rate and usual incidence of hospital stays. Unfortunately, no compiled database with the total number of beds activated during the epidemic wave has been made publicly available to check this hypothesis. Nevertheless, the absence of significant associations between basal population health and healthcare services, and the incidence and mortality rates reinforces the assumption of an important hospital adaptation to the epidemic wave.

As expected (4), in-hospital mortality and lethality rates were significantly higher in departments with older population. Age has indeed been identified as the main risk factor of disease severity and death in repeated cohort studies (15,32–35). We did not include prevalence of comorbidities such as diabetes or obesity in our study, as available data at department level are scarce. But we considered several indicators associated with overall population health such as basal mortality rate and the usual incidence rates of hospital stays in endocrinology, cardiology, pneumology and medicine wards.

We found no significant association between incidence of new chloroquine and hydroxychloroquine (CQ/HCQ) dispensations in pharmacies and in-hospital COVID-19 incidence, mortality or lethality at department level. We could not include in-hospital dispensations data, which are not publicly available in France, but the CQ/HCQ prescription for COVID-19 was limited by French regulation on May 26^th^, 2020, after the end of our study period. Interestingly, although univariate analyses showed important and significant associations between in-hospital incidence or mortality rates and new CQ/HCQ dispensations, these associations were positive (the more CQ/HCQ dispensations, the higher incidence or mortality rates). This suggests a common confounding by indication (55), which was corrected by the multivariate analysis. We included CQ/HCQ dispensations in our models, as their responsibility in the lower COVID-19 burden observed in some regions has been mentioned. But our results do not support this hypothesis, as CQ/HCQ dispensations were not significantly associated with in-hospital incidence, mortality or lethality at the department level. Besides, our multivariate analysis explained the near totality of in-hospital COVID-19 incidence in Paris and Bouches-du-Rhône departments, the two departments where CQ/HCQ dispensations were highest. The observed mortality in Bouches-du-Rhône was even higher than predicted by our model. Conversely, lethality was slightly overestimated in this department. Nevertheless, our ecological study was not designed to provide any arguments for or against the effectiveness of CQ/HCQ against SARS-CoV2 at individual level (56–61).

Our multivariate model, adjusted on spatial heterogeneity, did not identify any significant association between in-hospital COVID-19 incidence and climate. This contrasts with previous reports (7–14), and may be explained by our choice to use climate characteristics rather than meteorological factors, such as temperature, humidity or rainfall. But this matches with the current summery explosion of the pandemic across the United States of America, Southern America as well the current second wave in European countries (15).

Similarly, our study did not exhibit any significant association between economic indicators and in-hospital COVID-19 incidence or mortality at the department level. This contrasts with a recently pre-published study that showed a larger impact of the pandemic on overall mortality in the poorest of the 35,000 French municipalities (62). The spatial scale of our analysis (department level) may have been too wide to identify such an association. The multidimensional reduction of economic indicators using HAC on PCA, which allows assessing numerous factors and takes into account collinearities and the curse of dimensionality, may also have flattened differences between departments and hidden possible associations.

Our study may also have suffered from several additional limitations. We only analysed COVID-19 in-hospital data. Testing was indeed too limited in France during the first epidemic wave to provide an accurate estimation of outpatient incidence. Besides, casualties in retirement homes have not yet been made available at department level. Hospitalization criteria may not be homogeneous across departments, and milder cases may have been hospitalized more frequently in less affected departments or depending on local care policy (63). Age of hospitalized patients have not been made available at department level to account for these differences. This may partly explain why our multivariate analysis explained only 53.1% of lethality, with a slight overestimation of the lethality rate in Bouches-du-Rhône. Additional factors may have been associated with lethality, needing further analysis at a more accurate scale. Finally, we performed an ecological study, not an individual population study, which introduces classical ecological fallacy, and therefore means we cannot infer any direct individual risk (64).

In conclusion, our study could explain a great part of the spatial heterogeneity of in-hospital COVID-19 incidence and mortality across metropolitan France, and to a lesser extent of its lethality. We highlighted that the population age structure was an important determinant of the mortality and lethality at the department level, and that the lockdown policy was an effective way to control the diffusion and the severity of the epidemic wave across the country. All three rates, in-hospital incidence, mortality and lethality, were strongly determined by the initial diffusion of the pandemic before the national lockdown froze the situation from March 17^th^, 2020. We recommend that ecological studies comparing COVID-19 morbidity or mortality between countries take into account important confounders such as differences in population age structure, testing capacities, the delay since the start of the epidemic heterogeneous, and mitigations strategies.

## Data Availability

All data is publicly accessible. Links to the source data are indicated in the manuscript.

https://www.data.gouv.fr/fr/datasets/donnees-hospitalieres-relatives-a-lepidemie-de-covid-19/

https://www.insee.fr/fr/statistiques/1893198

https://drees.solidarites-sante.gouv.fr/etudes-et-statistiques/publications/article/nombre-de-lits-de-reanimation-de-soins-intensifs-et-de-soins-continus-en-france

https://www.ansm.sante.fr/S-informer/Points-d-information-Points-d-information/Usage-des-medicaments-en-ville-durant-l-epidemie-de-Covid-19-point-de-situation-apres-cinq-semaines-de-confinement-Point-d-information

https://www.insee.fr/fr/statistiques/4487988?sommaire=4487854

https://www.insee.fr/fr/statistiques

https://cartographie.atih.sante.fr

https://www.data.gouv.fr/fr/datasets/admin-express/

## Notes

### Competing Interest Statement

The authors have declared no competing interest.

### Clinical Trial

This study is an ecological analysis, using anonymous and aggregated, publicly available data.

### Funding Statement

no external funding are declared

### Author Declarations

This study is an ecological analysis, using anonymous, aggregated, publicly available data. According to the regulations, the agreement of an ethics committee is not required in this context.

